# Multilevel estimation of the relative impacts of social determinants on income-related health inequalities in urban Canada: Protocol for the Canadian Social Determinants Urban Laboratory

**DOI:** 10.1101/2024.12.31.24319827

**Authors:** Charles Plante, Suvadra Datta Gupta, Thilina Bandara, Daniel Beland, Olivier Bellefleur, Christine Blaser, Cheryl Camillo, Eileen de Villa, Daniel Dutton, Daniel Fuller, Jasmine Hasselback, Lisa Marie Lix, Anousheh Marouzi, Nazeem Muhajarine, Geranda Notten, Bill Reimer, Michael Wolfson, Marisa Young, Daniel Yupanqui Concha, Cory Neudorf

**Author notes:** **<u>About the Urban Public Health Network (UPHN) Research Group:</u>** The Urban Public Health Network (UPHN), a national organization founded in 2004, comprises the Medical Officers of Health from 25 of Canada’s major urban centers. Working collaboratively and with a collective voice, the network addresses public health issues that are common to urban populations. The UPHN Research Group, a coalition of independent researchers and students, supports these efforts by generating and mobilizing relevant evidence. Research operations for the UPHN are conducted in partnership with the University of Saskatchewan. **Suggested Citation:** Plante Charles, Datta Gupta Suvadra, Bandara Thilina, Beland Daniel, Bellefleur Olivier, Blaser Christine, Camillo Cheryl, De Villa Eileen, Dutton Daniel, Fuller Daniel, Hasselback Jasmine, Lix M. Lisa, Marouzi Anousheh, Muhajarine Nazeem, Neudorf Cory, Notten Geranda, Reimer Bill, Wolfson Michael, Young Marisa, Yupanqui Concha Daniel, and Neudorf Cory. 2024. “Multilevel estimation of the relative impacts of social determinants on income-related health inequalities in urban Canada: Protocol for the Canadian Social Determinants Urban Laboratory”. MedRxiv.

## Abstract

Building on Canadian data at the provincial, regional, community, and personal levels, the Canadian Social Determinants Urban Laboratory (CSDUL) will enable multilevel and longitudinal investigation of how social determinants of health (SDOH) impact population health (both mental and physical) and health inequities in Canada. Utilizing administrative data linkage, CSDUL will be developed by combining social, economic, and political mechanisms at multiple levels, from national to individual, following the World Health Organization (WHO) SDOH framework. Organized using a hub-and-node model, CSDUL will be created by validating unit and area-level indicators and merging survey and administrative data to provide a comprehensive understanding of SDOH at micro, meso, and macro levels. The project will replicate WHO/Europe’s decomposition analysis of income-related inequalities in self-reported health, assessing the relative impact of social determinants on health outcomes.

**Extended Abstract:** *Introduction:* Two decades of research have highlighted persistent income-related health inequities in Canada at municipal, provincial, and national levels. This project aims to examine how social, economic, and political factors create conditions that shape health inequalities, and investigate how structural and intermediate determinants explain health disparities across national, provincial, city, neighbourhood, and individual levels.

*Methods:* We will create the Canadian Social Determinants Urban Laboratory (CSDUL), a multilevel, longitudinal virtual environment combining multiple surveys and administrative databases, guided by the WHO Social Determinants of Health framework. Initially covering 2011-2015, CSDUL will expand as more data becomes available. Organized in a hub-and-node model, it will include a central hub and five project nodes. We will develop and validate area-based indicators, merged with data to provide a comprehensive understanding of social determinants of health at micro, meso, and macro levels^1^.

*Results:* The primary research deliverables of this project will be to critically analyze the strengths and limitations of survey and administrative databases for health research and develop methods for deriving variables from them. After developing CSDUL, we will replicate WHO/Europe’s income-related health inequality analysis for urban Canada and report on the impact of social determinants on health outcomes.

*Discussion:* A key strength of the proposed virtual data laboratory is its ability to examine how various determinants affect health at different levels and explore their impact on identifiable groups (e.g., by gender). It highlights the multifactorial nature of health and identifies the factors most likely to drive health outcomes, such as what makes Canadians healthy or sick.

*Conclusion:* Multisectoral interventions are most effective when they are customized to meet the unique needs of specific sub-populations, using robust and multilevel data sources like CSDUL.

**Key Messages:**

- Building on Canadian data, the Canadian Social Determinants Urban Laboratory (CSDUL) is the first initiative of its kind to provide a comprehensive understanding of how social determinants impact health outcomes in Canadian cities.
- CSDUL will be a multilevel, longitudinal data Laboratory, organized using a hub-and-node model, operationalizing the WHO Social Determinants of Health framework
- After developing CSDUL, we will replicate WHO/Europe’s decomposition analysis of income-related inequalities in self-reported health for urban Canada

## Introduction

Despite a mounting consensus among researchers, advocates, and health professionals that social determinants are the primary driver of the health of Canadians, the empirical basis for this claim remains thin. Studies from several European countries have shown that income and social security are primary drivers, with social determinants of health explaining up to 89% of the variation in self-reported health in these countries (World Health Organization 2019). A similar trend is observed in the United States, where socioeconomic factors tend to be the strongest predictors of health outcomes (Hood et al. 2016). One of the very few flagship reports describing health disparities across Canadian cities, Reducing the Gaps in Health: A Focus on Socio-Economic Status in Canada, championed by the Urban Public Health Network (UPHN), found that income-related health inequalities have remained largely unchanged since the early 2000s and vary considerably between cities and indicators (Public Health Agency of Canada (PHAC) 2018; Plante C, Missiuna S and Neudorf C 2021; Missiuna et al. 2021). Many of these inequalities persist at municipal, provincial, and national levels and are direct consequences of individuals’ socio-economic disadvantages (Public Health Agency of Canada (PHAC) 2018; CIHI 2016). As social determinants of health are many, and their interactions are complex (Council of Canadian Academies 2015; Braveman and Gottlieb 2014), a more in-depth account of how social determinants impact health is needed to identify the highest-impact targets for interventions that can reduce these inequalities and improve health outcomes.

Considerable data is required to measure different aspects of the social determinants of health, and this data has to be combined, motivating calls for more significant investments in linkages between health and social data (Public Health Agency of Canada (PHAC) 2022, 2021b; Dusetzina et al. 2014; Bradley et al. 2010). During the past two decades, Canadian research data access centers have developed a new way of conducting health and social research using nationally representative linked surveys and administrative data (Currie and Fortin 2015). However, despite offering a significant amount of data on the various social determinants of health, the advanced knowledge of these data sources that is needed to work with them is limited among researchers and academics (Canadian Research Data Centre Network, n.d.). Considerable disparities also exist in the capacity of local health units to survey regional differences in population health and health equity, with some units having no capacity at all (Potvin 2014; Guyon and Perreault 2016) and becoming familiar with large and complex data sources like those in the RDC is costly. Even among units that do have the capacity to make these investments, work tends to be carried out in isolation, and no national standards exist to ensure that research is carried out in ways that support comparative learning (Public Health Agency of Canada (PHAC) 2021a; Wilson et al. 2017; Canadian Institute for Health Information (CIHI) 2016).

This paper presents an innovative protocol for the world’s first fully integrated virtual, multilevel, and longitudinal social determinants laboratory environment entitled Canadian Social Determinants Urban Laboratory (CSDUL). CSDUL aims to facilitate multi-level statistical analyses of how social and structural determinants of health impact the overall health of Canadians-achieved by creating a set of programs, algorithms (statistical codes/syntaxes), and data components accumulating information from at least 15 primary data sources within the secure research environment of the RDC.

The project will further use CSDUL to replicate the decomposition analysis^2^ of income-related inequalities in self-reported health carried out by the WHO/Europe (Brown and Yang 2019) for urban Canada. In short, this project will allow us to determine how much of the variation in income-related health inequalities in Canada can be attributed to the various social determinants of health. i.e., report on the relative impact of social determinants on health inequities. In addition to replicating past studies that have illustrated the contribution of social determinants of health (SDOH) in determining health status, the CSDUL environment will allow us to examine how different categories of SDOH operate on health at different levels and bridge the gap between the WHO’s conceptual and action frameworks.

## Methods

### Data

The backbone of CSDUL’s data structure will be Statistics Canada’s Canadian Population Health Survey (CPHS), which researchers can access via Research Data Centre (RDC) programs. The CPHS consists of linked survey, administrative, and registry data among the Canadian Community Health Survey (CCHS), the Discharge Abstract Database (DAD), the National Ambulatory Care Reporting System (NACRS), the Canadian Vital Statistics Death Database (CVSD), the Canadian Cancer Registry (CCR), and 20 years of Canada Revenue Agency (CRA) personal and familial tax records. These data will be merged with area-level indicators using historical postal codes in the CPHS, Canada Post’s Postal Code Conversion File Plus (PCCF+), and Census Boundary Files provided by Statistics Canada. Additional data sources include the Canadian Classification of Functions of Government (CCOFOG), data from the Canadian Urban Environmental Health Research Consortium (CANUE), DMTI Enhanced Points of Interest (DMTI-EPOI), and Statistics Canada’s Social Environment Typologies (CanSET). Table 1 provides a brief description of the datasets. The time period with existing data for all these sources is 2011-2017. This will be the initial observation window for CSDUL, but we aim to extend it in future studies as new data becomes available. The CCHS samples approximately 65,000 Canadians each year, and we will add new years as they become available, resulting in an overall *N* of approximately half of one million individual Canadians and a number of health events (e.g. hospitalizations or deaths) that is an order of magnitude greater.

**Table 1.**
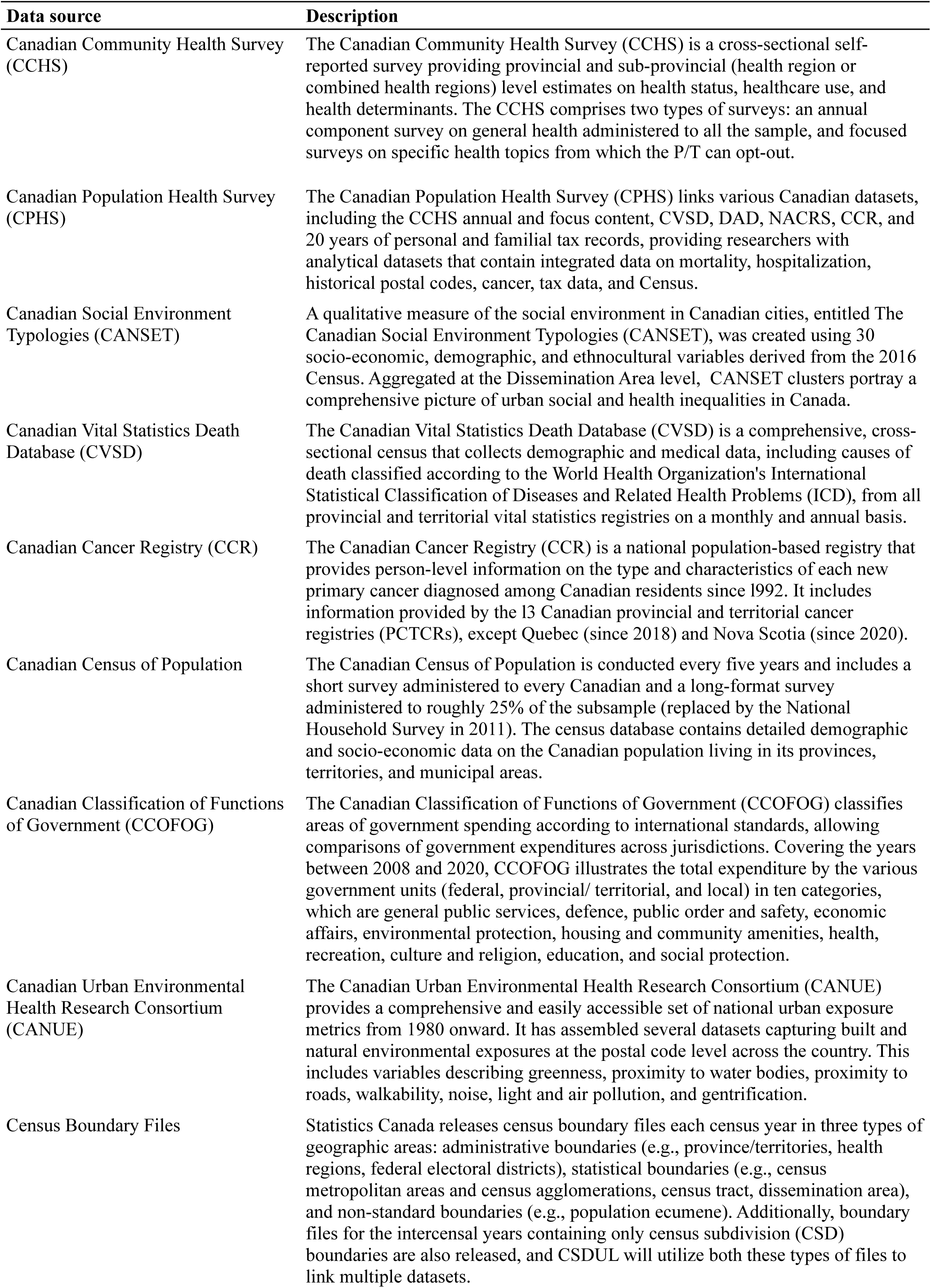
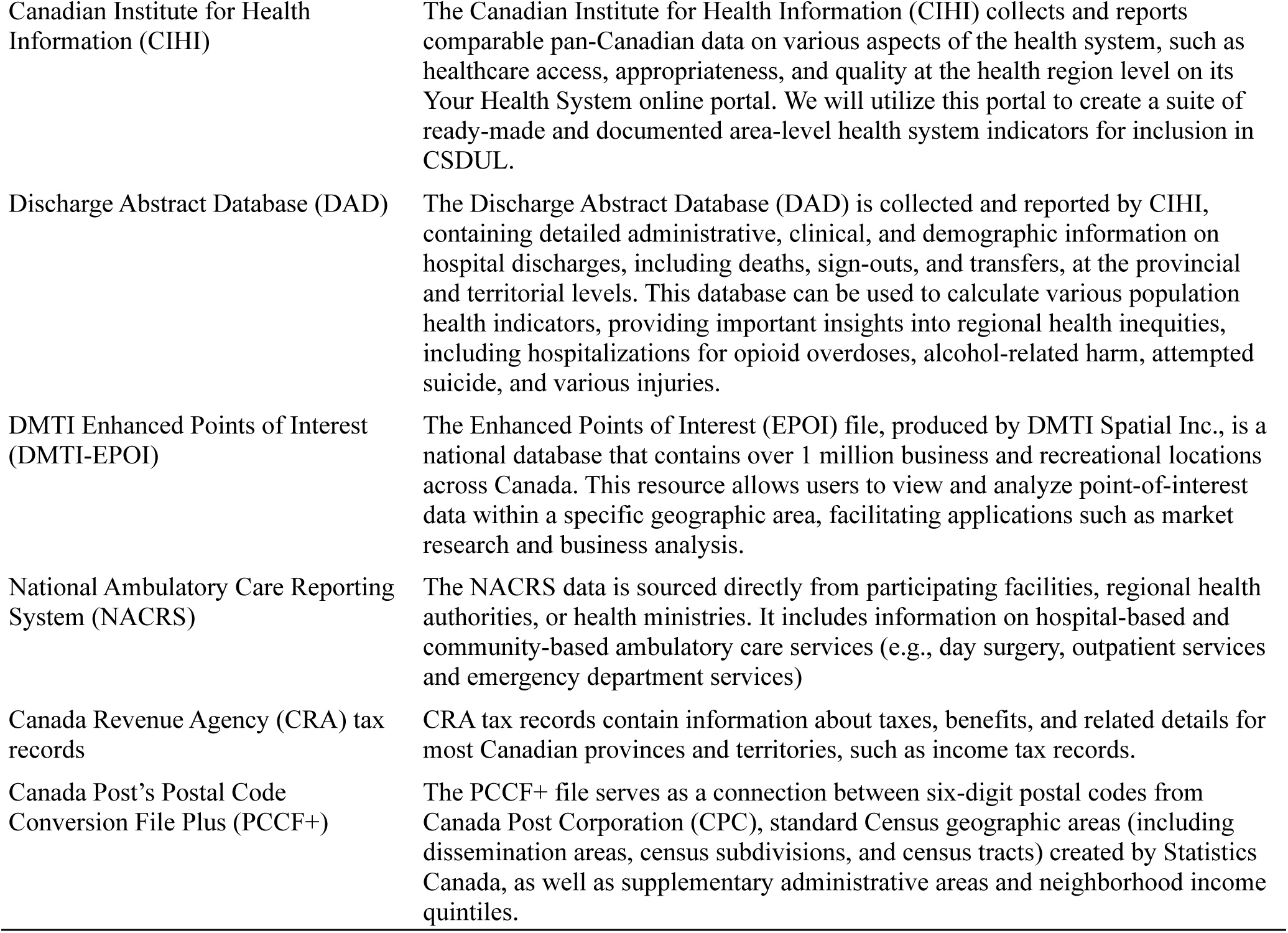
Description of the datasets.

Furthermore, at this time, we will restrict CSDUL to Canada’s cities and towns because our aim is to estimate a fully articulated model of the social determinants of health, and these factors are not as well understood or measured in rural and remote regions (CIHI 2006; Lavergne and Kephart 2012; Pampalon, Hamel, and Gamache 2010). We will classify communities as cities and towns if they qualify as census agglomerations or census metropolitan areas—that is, communities with a core population of at least 10,000 (Statistics Canada 2017a); these entities can be made up of multiple municipalities (e.g., the census metropolitan area of Montreal includes Laval among others)—this was the same approach used by Statistics Canada in creating CanSET (Subedi, Aitken, and Greenberg 2022).

In their raw form, most of the databases that will be included in CSDUL are not user-friendly and not readily usable for health analysis. Thus, many of the primary research deliverables of this project will be to critically analyze and document the strengths and limitations of these databases for supporting health research and create and validate the best methods for deriving variables from them. By combining all these data elements, our project will create a multilevel and longitudinal virtual laboratory environment that can be used to examine the operation of social determinants on health at micro, meso, and macro levels and to study income-related health inequalities. This approach builds on previous studies that could only demonstrate the contributions of SDOH to health across a limited subset of the WHO SDOH framework elements (Hood et al. 2016; Park et al. 2015).

The development of CSDUL will be organized using a Hub and multidisciplinary Node teams across Canada and organized around the WHO Social Determinants of Health framework. Figure 1 provides an overview of the CSDUL project and its mappings between data sources and Hub and Node activities. Figure 2 provides an overview of the planned construction of the CSDUL environment, its elements, and its variables.

**Figure 1.**
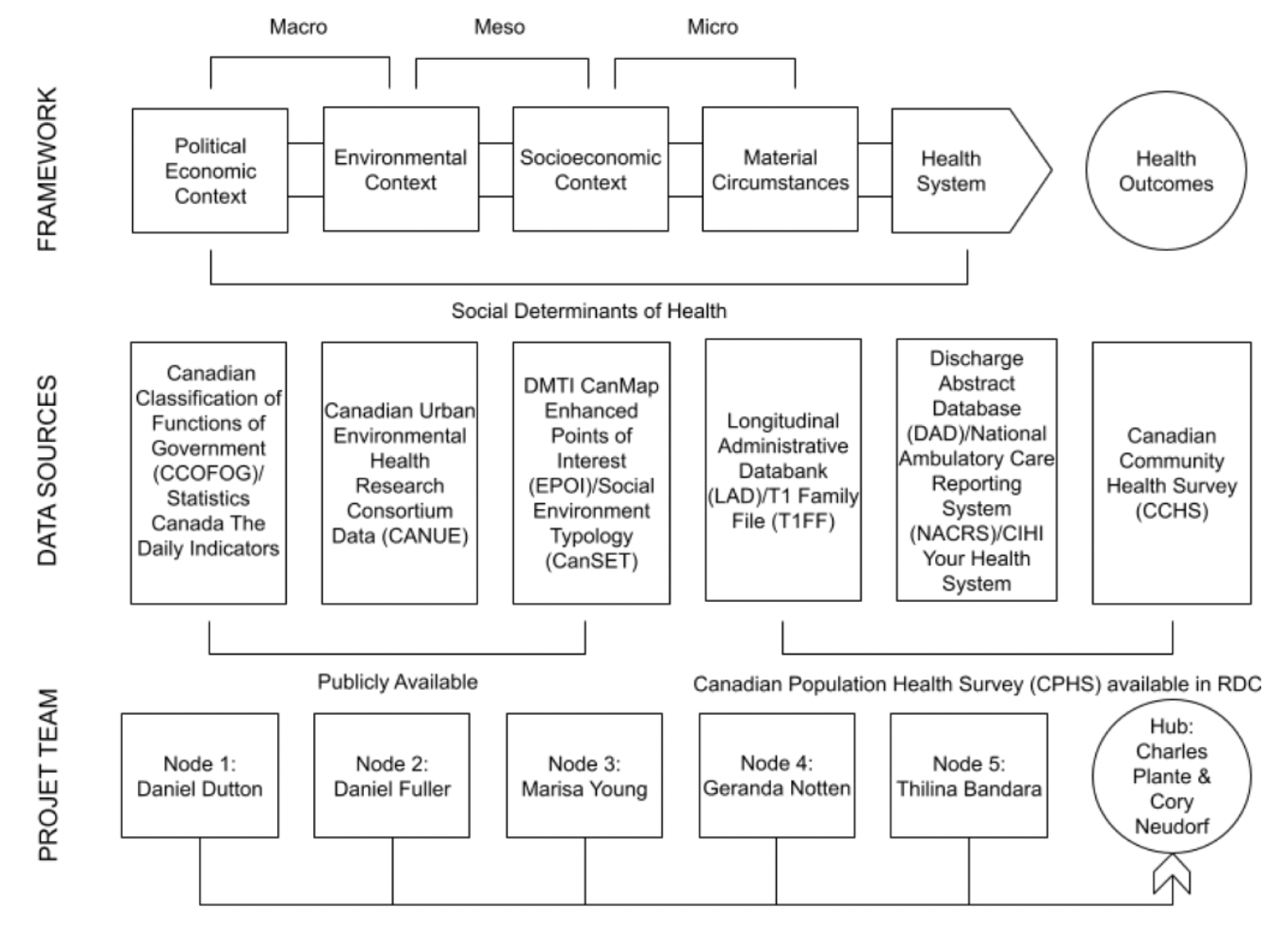
Stylized overview of Canadian Social Determinants Urban Laboratory (CSDUL), its data structure, and its organizing Nodes. Notes: RDC: Research Data Centre.

**Figure 2.**
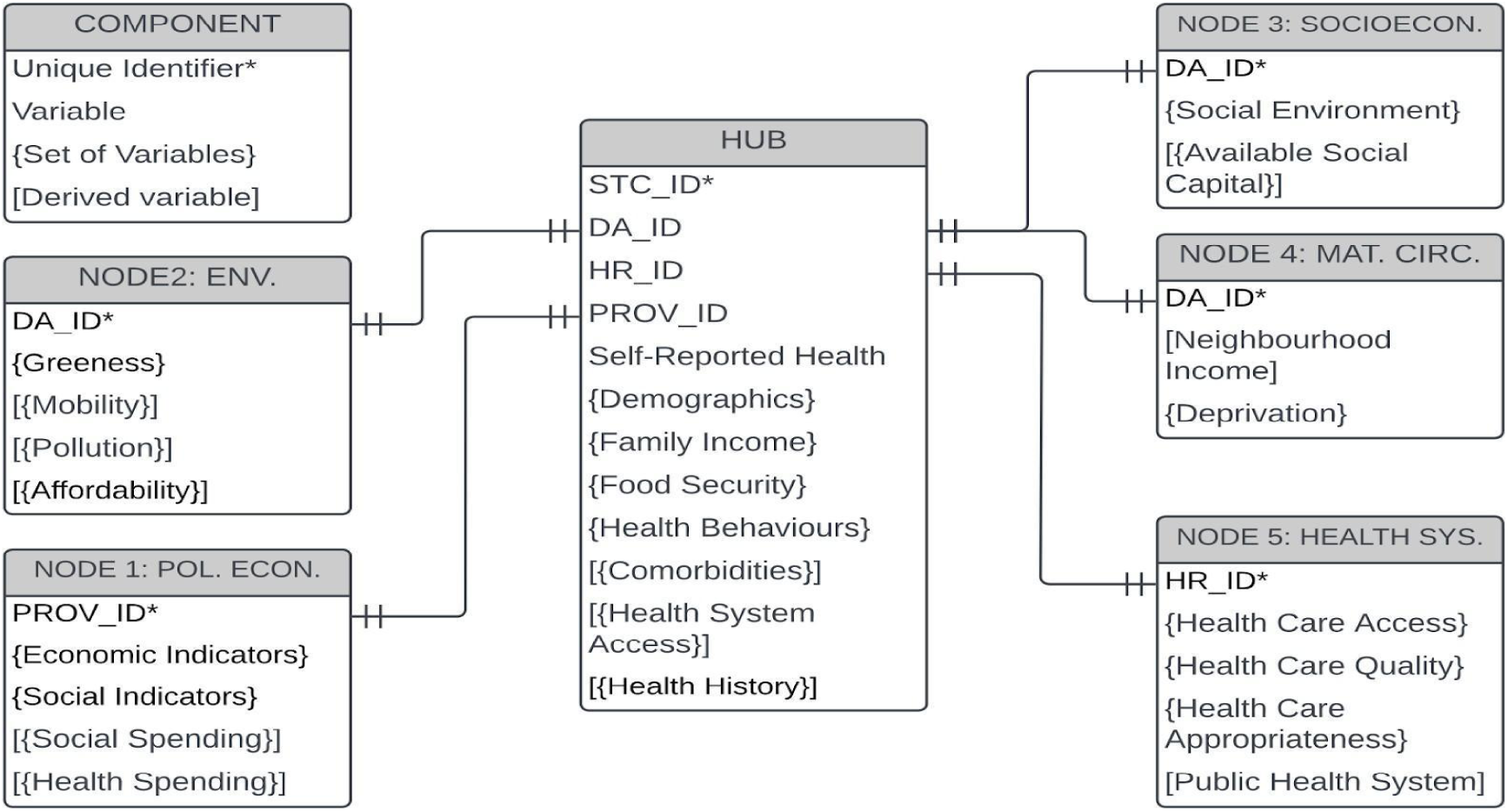
Description of the Canadian Social Determinants Urban Laboratory (CSDUL) data components and their relationships. Notes: DA_ID: Dissemination Area Identification Number. PROV_ID: Province Identification Number, STC_ID: Survey Respondent Identification Number. HR_ID: Health Region Identification Number. Env.: Environmental Context. POL. ECON.: Political Economic Context. SOCIOECON.: Socioeconomic Context. MAT. CIRC.: Material Circumstances. HEALTH SYS.: Health System.

**Figure 3.**
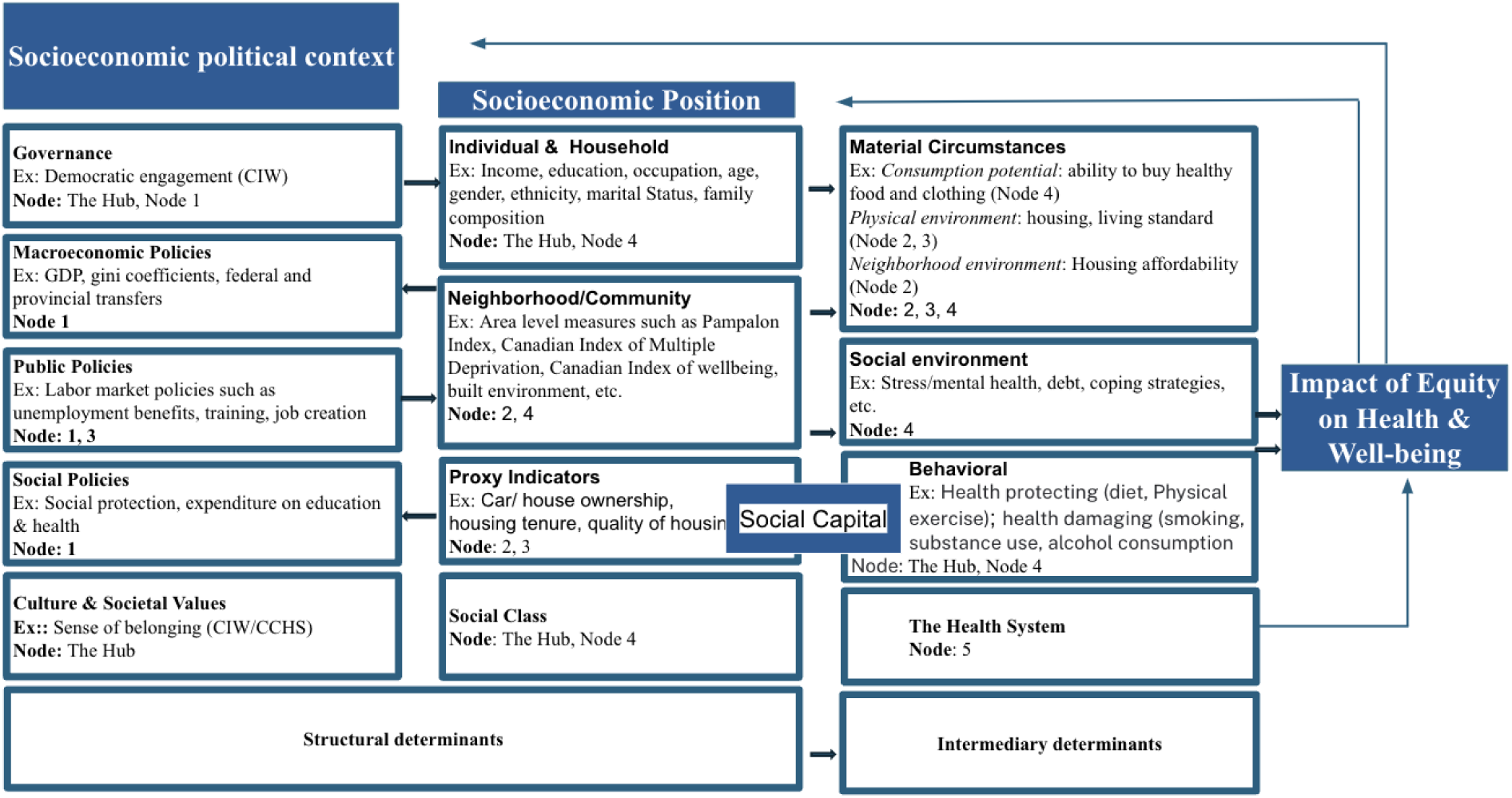
Operationalizing the WHO Social Determinants of Health Framework. Notes: 1. CIW: Canadian Index of Wellbeing 2. This framework is an adapted version of the WHO SDOH framework, with variables and Nodes mapped to each of its components. However, it does not provide a complete list of the variables that will be generated by the Nodes for use in our multivariate analysis.

#### Outcome variable

Self-reported health will be the primary outcome variable of interest for the decomposition analysis. Drawing on validation work by our team using the Health Utility Index (Horsman et al. 2003)(Plante, Missiuna, and Neudorf 2024), we will categorize self-reported health based on whether respondents reported having health that is at least better than “fair.” Self-rated health is a widely used indicator of overall health status, and multiple studies have validated its usefulness as a subjective proxy for a number of objective health measures (Schnittker and Bacak 2014; Wu et al. 2013; Falconer and Quesnel-Vallée 2017; Vaillant and Wolff 2012).

#### Predictor variables

Each Node will contribute tables of predictor variables to the decomposition analysis based on their work on developing CSDUL. To achieve this, working group Nodes will conceptualize, operationalize, and link constructs from the WHO conceptual and action social determinants frameworks. Hub individual-level variables will include demographic variables (e.g., gender and age), family income, health behaviours (Canada 2018), health comorbidities (Glasheen et al. 2019; Quan et al. 2005), health systems access (Canadian Institute for Health Information (CIHI), n.d.-b), and health history (Le Meur, Gao, and Bayat 2015; Nguena Nguefack et al. 2020). Political and economic context variables will include economic indicators (e.g. macroeconomic policies, gini coefficient, etc.) social indicators (e.g. policies on social welfare, land and housing, etc.) social spending (Dutton et al. 2018), and health spending (Dutton et al. 2018) at the health region and provincial level. Environmental context variables will include greenspace, (Crouse et al. 2021) walkability (Herrmann et al. 2019), gentrification (Firth et al. 2021), urban sprawl (Luan and Fuller 2022), and air pollution (Weichenthal et al. 2021) at the dissemination area level in the urban areas. Socioeconomic context variables will include the social environment such as stress and different types of available social capital, including what Klinenberg refers to as social infrastructure (Klinenberg 2018). Material circumstances will include area-based income and deprivation. Health systems indicators will include health care access, health care quality, health care appropriateness, and public health systems.

### CSDUL development

A central hub operating out of the Urban Public Health Network Research Group and based at the University of Saskatchewan (hereinafter referred to as the Hub) will assemble data in collaboration with partners, combining them with individual-level indicators of health behaviour, history, system access, system utilization and outcomes in the CPHS. The Hub will also coordinate the project and ensure the primary objectives are met. Under the Hub, CSDUL will have five project nodes, led by different investigators, who will create and validate area-based indicators at macro and meso levels that will be integrated with CSDUL and the decomposition analysis (objective 2). The Nodes are-Political and Economic Context (Node 1), Environmental Context (Node 2), Socioeconomic Context (Node 3), Material Circumstances (Node 4), and Health Systems (Node 5). Additionally, the Hub will assemble and disseminate this virtual environment, which will operate on individual-level data within Statistics Canada’s Research Data Centers. The project will be advised by a Senior Advisory Group (SAG), composed of experts in the field of population health and secondary data development and use. Additionally, CSDUL will be advised by a Knowledge Users Group, operating as part of the Urban Public Health Network’s (UPHN) unique Integrated Knowledge Translation (IKT) Collaborative (Malkin et al. 2021), which brings together academic researchers, national data stewards, and local public health practitioners eager to advance measures and tools that can be used for effective local-level evidence-based decision-making.

#### Node 1. Political and economic context

The primary objective of this node will be to study and document the association between macroeconomic factors and government policy on health outcomes using the new Canadian Classification of Functions of Government (CCOfOG) data (Statistics Canada 2023a). Statistics Canada has recently updated its Consolidated Government Revenue and Expenditures tables to align with international government revenue and spending practices. This update brings a significant amount of detail, particularly in the areas of health and social spending. This node will create a simplified version of this data (spanning from 2008 to 2018) and a set of guidelines for CSDUL. Several macro-level variables from Statistics Canada’s The Daily Indicators series, e.g., Gini coefficients, GDP, and the unemployment rate, will be included.

#### Node 2. Environmental context

Node 2 aims at measuring the impacts of environmental factors, i.e., walkability, bikeability, food environments, transit accessibility, gentrification, and housing affordability, on health outcomes in Canada’s cities since 2010. The measures already developed are walkability, urban sprawl, bikeability, food environments, transit accessibility, and gentrification. It will also create new neighborhood-level indicators of environmental context using the CANUE data specifically relating to housing affordability, as well as validate these measures. CSDUL will be one of the 1st large-scale validation studies explaining how environmental factors can be combined in population health research with population health outcomes, including self-reported health.

#### Node 3. Socioeconomic context

Node 3 will expand on the existing theoretical and methodological work conducted on the area-based available social capital in rural communities in Canada (W. Reimer 2004; B. Reimer 2011), classifying it into four normative types: market, bureaucratic, communal, and associative for urban Canada. This work will be expanded by integrating recent approaches to capturing the social infrastructure in residential areas (Klinenberg 2018; Young et al. 2023; Young & Singh 2022) This Node will also make use of the UPHN work with the Health Analysis Division of Statistics Canada, which produced a new “qualitative” measure of the social environment-the Canadian Social Environment Typology (CanSET) (Subedi, Aitken, and Greenberg 2002). The key purpose of Node 3 is to investigate the connection between the availability of family-friendly resources from the Digital Mapping Technologies Inc. (DMTI) Spatial (see (Young, Leipe, and Singh 2023; Young and Singh 2024) for an overview). Node 3 will also use leading indicators of population health in Canadian cities, starting from 2010 from DMTI and other sources.

#### Node 4. Material circumstances

The primary objectives of Node 4 will be to develop and support the distribution of improved dissemination area-level median after-tax income measures for the CSDUL project and the decomposition analysis. In Canada, income-related health inequalities at the area level have traditionally been measured using income data collected once a year by the Census. However, in recent times, social scientists have preferred to use median after-tax income as a more accurate measure of disposable income (Murphy, Zhang, and Dionne 2010; Brady 2003). Canada also has an Longitudinal Administrative Databank (LAD) that provides longitudinal data on income at the dissemination area level since 1982, but it has not been widely used in health science research. Node 4 extends the utilization and distribution of a new after-tax income area-level indicator based on the LAD. It aims to assess the consequences of implementing a new indicator to explore health inequalities associated with income and to study how our preference for area-level income or deprivation impacts our research on health inequalities.

#### Node 5. Health system

Node 5 research activities will focus on developing and deploying a suite of area-level (primarily at the health region level) health system indicators to quantify system-level access and quality of care that can be integrated with CSDUL (Plante et al. 2021; Malkin et al. 2021). This will involve deriving health system indicators that are available at the sub-provincial level and identifying and assessing individual-level indicators of health system access and use from CCHS.

#### Senior Advisory Group and Knowledge Users Group

The Knowledge Users Group and the Senior Advisory Group, integral parts of the UPHN IKT Collaborative around the CSDUL project, are composed of the UPHN membership, which includes the country’s top public health medical leadership and the country’s top experts in population health research, respectively. These two groups will receive regular updates from the Hub and will provide valuable advice on the design and dissemination of CSDUL relating to their areas of expertise and advanced experience. They will also play a key role in facilitating knowledge mobilization within their organizations and among their affiliate health leadership agencies.

### Decomposition Analysis

Our study will follow the WHO/Europe(World Health Organization (WHO) 2019) and use a “twofold” Oaxaca-Blinder decomposition (Jann 2008; Neumark 1988; Oaxaca and Ransom 1994) to identify the primary drivers of self-reported health. We are mindful that the CCHS has been redesigned multiple times (most recently in 2015); we will follow best practices(Thomas and Wannell 2009) and only work with variables that have been measured in the same way over time as we have done in previous work.(Plante, Missiuna, and Neudorf 2021) As was done by WHO/Europe, we will also report on the distribution of social determinants and health outcomes in the cities and towns we include and replicate our analysis for additional self-rated outcomes: mental health and life satisfaction.

The decomposition analysis will be done as follows. If y_q_ is a vector of observations of dichotomized self-reported health in quintile q, and X_q_ a matrix of observed social determinants and a constant, then the difference between average self-reported health in the lowest quintile, ȳ_1_, and the highest quintile, ȳ_5_, can be fitted using linear regression and decomposed into two components. First, a component that is attributable to differences in X_q_ between the quintiles (i.e., differences in their composition) and, second, a component that is attributable to differences in associations between y_q_ and X_q_, captured by the coefficients in the linear model, β_q_. If we use the associations of the fifth and richest quintile, β_5_, as our benchmark for “best health,” then the estimated difference in average self-reported health between that quintile and the worst-off quintile, β_1_, is decomposable as:

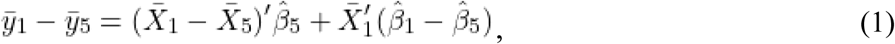

If social determinants in X_q_ are indexed by m, then the estimated proportional contribution of determinants m ∈ G is:

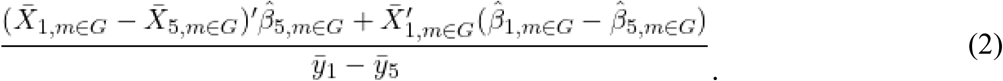

We will estimate Equation (2) for each group of categories of social determinants and levels. Our baseline estimates of β_q_ will use pooled multilevel logistic regression with mixed effects and include all predictors in X_q_ identified by our Nodes (use of logistic regression will slightly modify the above notation, but the interpretation will remain the same (Bauer and Sinning 2008; Sinning, Hahn, and Bauer 2008)).

Data will be pooled to ensure sufficiently large local sample sizes (in past work,(Plante, Missiuna, and Neudorf 2021) we have found pooling five or more years is needed—at this time, we only have seven years; not enough for two five-year pooled intervals). We will include indicators for year to control for period effects. Provincial effects will be set to fixed, and meso-level variable effects (i.e., health region, metro-area or municipality), random. As this is the first study of its kind, we will explore different model specifications and document the implications of these choices for our decomposition results (Gomila 2021; Hellevik 2009).

All statistical estimates will be calculated using probability weight, and standard errors will be calculated using bootstrapping to account for the complex survey design of the CCHS (Rao 2006).

### Interaction/Effect Modification Analysis

We will also examine the impacts of accommodating interaction/effect modification (Corraini et al. 2017) between certain predictors—specifically, self-reported gender (i.e., male, female, and, when possible, non-binary) and different categories of non-white identifying (e.g., black identifying)—and fitting random effects at lower levels (e.g., aggregate dissemination area or census tract). This would allow us to see, for instance, if the impact of education is more significant for women than men (Hill and King 1995; Everett, Rehkopf, and Rogers 2013).

## Results

In the beginning year of the project, the Hub will take primary responsibility for integrating the work and components completed by the Nodes in the RDC and creating and disseminating CSDUL. It will also work with the nodes simultaneously to create and validate social and environmental area-based indicators at macro and meso levels that will be merged with the survey and administrative data. It will also construct individual-level demographic variables, health behaviour indicators, and health history/health system use indicators. Individual health behaviours (i.e., smoking, drinking, physical activity, and frequency of fruit and vegetable consumption) will be operationalized in the CCHS using established methods developed by Statistics Canada (Canada 2018). Demographic variables, including age, gender, ethnicity, marital status, family composition, level of education, family income, employment status, and occupation, will be added. For health history/health system use, DAD and NACRS data combined with International Classification of Diseases (ICD) coding will be used to identify comorbidities (i.e., the Charlson Comorbidity Index). In the second and third years of the project, the Hub will complete the decomposition of the income-related inequalities of self-reported health.

## Discussion

The World Health Organization (WHO) Commission on the Social Determinants of Health (CSDOH) has called for a “third wave” in population health research, which “makes explicit that health systems and the people who use them exist within a social context that can powerfully determine people’s chances to be healthy—not only through access to health services, but also through access to a range of other resources, opportunities, and rights’ (Östlin et al. 2011). There are numerous social determinants that can impact health outcomes, and the way these factors interact with each other is complex (Braveman and Gottlieb 2014; Mikkonen and Raphael 2010). For example, political and economic context refers to a broad set of structural and functional aspects of the society that cannot be directly measured at the individual level but have an immense influence on the patterns of social stratification and, therefore, on the health of populations (World Health Organization (WHO) 2010). The environmental conditions in which populations live, work, and play have a profound influence on their health and well-being (World Health Organization (WHO) 2010). Exposure to harmful elements of the built environment—pollution, low-quality housing, and low amounts of green space, for example—can differ within cities and across subpopulations, leading to environmentally-mediated health disparities in specific conditions and overall health status (Gelormino et al. 2015). Biological factors, which include age and sex, also need to be considered when examining the impact of social determinants of health in health inequality research (World Health Organization (WHO) 2010). Material circumstances are the direct factors that determine an individual’s ability to live a healthy life (World Health Organization (WHO) 2010). These factors include income, education, employment status, and food security, among others. These factors are proximate measures of socioeconomic position that provide insight into the level of resources available to individuals to enable health-promoting conditions and behaviours (World Health Organization (WHO) 2010). In contrast, contextual determinants, such as the socioeconomic environment, provide individuals with the opportunity to live a healthy life (World Health Organization (WHO) 2010). The health system is considered an intermediary social determinant of health, and improving equity in health care requires addressing not only access but also the appropriateness and acceptability of care. Promoting intersectoral action to enhance these aspects can improve overall health outcomes and reduce differential quality of care (World Health Organization (WHO) 2010; Canadian Institute for Health Information (CIHI) 2013). The WHO SDOH framework has evolved, drawing on various models developed for different purposes, including public education and advocacy, and to better understand the complex interrelationships between social factors. It is important to note that these models, including the CSDOH referenced, have limitations. For instance, they often fail to account for critical aspects emphasized in Indigenous communities, such as the roles of culture and land. Our work aims to contribute to the ongoing evolution of the SDOH frameworks, offering a more comprehensive and inclusive perspective that reflects current research.

CSDUL will be the first of its kind that will bridge the gap between WHO’s conceptual and action frameworks in a data-driven way and clarify our understanding of what are the factors most likely to drive health outcomes in Canada, i.e., what makes Canadians sick or healthy. We will operationalize the WHO CSDOH framework, which considers the health system as a social determinant. Additionally, by linking multiple leading databases that provide information on the micro-meso-and macro levels, CSDUL will also help researchers to see how the various categories of this determinant operate on health at different levels, as conceptualized by WHO’s separate but complementary Framework for Action on Tackling Social Determinants of Health. It will also allow us to explore interaction effects with identifiable groups (including by gender) to understand how social determinants impact their health differently. Moreover, the data and decomposition methods employed by our study will allow us to combine and/or differentiate among associations between social and structural determinants and more effectively link their conceptualization with the levels at which decisions are made, and actions are taken.

CSDUL will also provide a framework and data components that can augment existing microsimulation tools like Statistics Canada’s Population Health Model (POHEM) (Hennessy et al. 2015), which are used to forecast the impacts of health policy decisions. Crucially, the development and future maintenance and dissemination of CSDUL will ensure that Canadian researchers are able to collaboratively use, build on, and learn from the work of others in the RDC. Researchers who are new to the RDC, including health system researchers, will no longer have to start from scratch, thus dramatically reducing the costs of doing population health and health equity analysis using Canada’s linked health and social data.

## Conclusion

Health inequities and inequalities are common in Canadian cities, but they are not equally distributed across various population groups. Due to data limitations, it has been difficult to conduct large-scale comparative research on health disparities in Canadian cities in the past. However, this situation is rapidly changing, and with the introduction of CSDUL, it will be possible to investigate social determinants of health at both individual and area levels simultaneously and seamlessly. Decomposition analysis is a method for understanding what contributes to health inequities by estimating associations between health inequalities, populations, and correlated determinants, such as those that our Hub and Nodes will identify, and analyzing the relative impact of each. Our work will go beyond the WHO/Europe analysis by providing data-driven relative contributions of individual-level variables, political and economic context, environmental context, socioeconomic context, material circumstances, and health systems.

## Data Availability

Not applicable

## Author Contributions

CP and CN conceived and designed the research protocol and secured funding. SDG and CP drafted the protocol manuscript, while members of the CSDUL team, including DF, DD, GN, MY, CB, BR, LL, CC, DB, TB, DY, and AM critically reviewed both the protocol and manuscript for methodological rigor and academic quality.

## Acknowledgments

The authors would like to thank Joanna Procyshen and Yvonne Hanson for their administrative support and the UPHN membership for their longstanding collaboration. Plante would also like to thank the Houston Family Trust for their early support. ChatGPT 3.5 was used as a writing aid. All views expressed in this work are our own.

## Ethics Declaration

The University of Saskatchewan Research Ethics Board deemed this study exempt (# E486).

## Footnotes

1 Macro refers to the national or federal level, meso to the provincial, city, and neighborhood levels, and micro to the individual or personal level.

2 Decomposition analysis is a method used to investigate changes in specific indicators by breaking them down into various contributing factors, enabling us to identify the underlying causes of these changes (Ang 2004).

## References

Ang, B. W. 2004. “Decomposition Analysis Applied to Energy.” In Encyclopedia of Energy, 761–69. Elsevier.

Bauer, Thomas K., and Mathias Sinning. 2008. “An Extension of the Blinder-Oaxaca Decomposition to Nonlinear Models.” AStA. Advances in Statistical Analysis. A Journal of the German Statistical Society 92:197–206.

Bradley, Cathy J., Lynne Penberthy, Kelly J. Devers, and Debra J. Holden. 2010. “Health Services Research and Data Linkages: Issues, Methods, and Directions for the Future.” Health Services Research 45 (5 Pt 2): 1468–88.

Brady, David. 2003. “Rethinking the Sociological Measurement of Poverty.” Social Forces; a Scientific Medium of Social Study and Interpretation. https://play.google.com/store/books/details?id=BuRBNQAACAAJ.

Braveman, Paula, and Laura Gottlieb. 2014. “The Social Determinants of Health: It’s Time to Consider the Causes of the Causes.” Public Health Reports 129 Suppl 2 (Suppl 2): 19–31.

Brook, Jeffrey R., Eleanor M. Setton, Evan Seed, Mahdi Shooshtari, Dany Doiron, and CANUE - The Canadian Urban Environmental Health Research Consortium. 2018. “The Canadian Urban Environmental Health Research Consortium - a Protocol for Building a National Environmental Exposure Data Platform for Integrated Analyses of Urban Form and Health.” BMC Public Health 18 (1): 114.

Brown, Chris, and Lin Yang. 2019. “Healthy, Prosperous Lives for All: The WHO European Health Equity Status Report Initiative.” Presented at the WHO Webinar.

Canada, Statistics. 2018. “Healthy Behaviours, 2017 - ARCHIVED.” Statistics Canada. June 26, 2018. https://www150.statcan.gc.ca/n1/en/catalogue/82-625-X201800154975.

Canadian Institute for Health Information. n.d. “Discharge Abstract Database (DAD) Metadata.” Accessed December 6, 2024. https://www.cihi.ca/en/discharge-abstract-database-dad-metadata.

Canadian Institute for Health Information (CIHI). 2013. “A Performance Measuremnt Framework for the Canadian Health Systems.”

Canadian Institute for Health Information (CIHI). 2016. “Pan-Canadian Dialogue to Advance the Measurement of Equity in Health Care: Proceedings Report,” July. https://secure.cihi.ca/estore/productFamily.htm?locale=en&pf=PFC3232&lang=en.

Canadian Institute for Health Information (CIHI). n.d.-a. “National Ambulatory Care Reporting System (NACRS) Metadata.” Canadian Institute for Health Information (CIHI). Accessed December 31, 2024. https://www.cihi.ca/en/national-ambulatory-care-reporting-system-nacrs-metadata.

Canadian Institute for Health Information (CIHI). n.d.-b. “Your Health System.” Canadian Institute for Health Information. Accessed May 3, 2024. https://yourhealthsystem.cihi.ca/hsp/indepth?lang=en#/.

Canadian Research Data Centre Network. n.d. “Strategic Plan 2019-2024.”

CIHI. 2006. “How Healthy Are Rural Canadians? An Assessment of Their Health Status and Health Determinants - a Component of the Initiative ‘Canada’s Rural Communities : Understanding Rural Health and Its Determinants.’” Canadian Institute for Health Information. https://play.google.com/store/books/details?id=0KJwDQEACAAJ.

CIHI. 2006. 2016. “Trends in Income-Related Health Inequalities in Canada.” Vol. 18. 10.12927/hcq.2016.24567.

Corraini, Priscila, Morten Olsen, Lars Pedersen, Olaf M. Dekkers, and Jan P. Vandenbroucke. 2017. “Effect Modification, Interaction and Mediation: An Overview of Theoretical Insights for Clinical Investigators.” Clinical Epidemiology 9 (June):331–38.

Council of Canadian Academies. 2015. Accessing Health and Health-Related Data in Canada: The Expert Panel on Timely Access to Health and Social Data for Health Research and Health System Innovation.

Crouse, Dan L., Lauren Pinault, Tanya Christidis, Eric Lavigne, Errol M. Thomson, and Paul J. Villeneuve. 2021. “Residential Greenness and Indicators of Stress and Mental Well-Being in a Canadian National-Level Survey.” Environmental Research 192 (January):110267.

Currie, Raymon F., and Sarah Fortin. 2015. Social Statistics Matter: A History of the Canadian RDC Network. Hamilton: Canadian Research Data Centres Network.

Dusetzina, Stacie B., Seth Tyree, Anne-Marie Meyer, Adrian Meyer, Laura Green, and William R. Carpenter. 2014. Linking Data for Health Services Research: A Framework and Instructional Guide. Rockville (MD): Agency for Healthcare Research and Quality (US).

Dutton, Daniel J., Pierre-Gerlier Forest, Ronald D. Kneebone, and Jennifer D. Zwicker. 2018. “Effect of Provincial Spending on Social Services and Health Care on Health Outcomes in Canada: An Observational Longitudinal Study.” CMAJ: Canadian Medical Association Journal = Journal de l’Association Medicale Canadienne 190 (3): E66–71.

Everett, Bethany G., David H. Rehkopf, and Richard G. Rogers. 2013. “The Nonlinear Relationship between Education and Mortality: An Examination of Cohort, Race/ethnic, and Gender Differences.” Population Research and Policy Review 32 (6): 893–917.

Falconer, James, and Amélie Quesnel-Vallée. 2017. “The Moderating Effect of Sociodemographic Factors on the Predictive Power of Self-Rated Health for Mortality in Canada.” Canadian Studies in Population [ARCHIVES] 44 (1-2): 77–99.

Firth, Caislin L., Benoit Thierry, Daniel Fuller, Meghan Winters, and Yan Kestens. 2021. “Gentrification, Urban Interventions and Equity (GENUINE): A Map-Based Gentrification Tool for Canadian Metropolitan Areas.” Health Reports / Statistics Canada, Canadian Centre for Health Information = Rapports Sur La Sante / Statistique Canada, Centre Canadien D’information Sur La Sante 32 (5): 15–28.

Gelormino, Elena, Giulia Melis, Cristina Marietta, and Giuseppe Costa. 2015. “From Built Environment to Health Inequalities: An Explanatory Framework Based on Evidence.” Preventive Medicine Reports 2 (September):737–45.

Glasheen, William P., Tristan Cordier, Rajiv Gumpina, Gil Haugh, Jared Davis, and Andrew Renda. 2019. “Charlson Comorbidity Index: ICD-9 Update and ICD-10 Translation.” American Health & Drug Benefits 12 (4): 188–97.

Gomila, Robin. 2021. “Logistic or Linear? Estimating Causal Effects of Experimental Treatments on Binary Outcomes Using Regression Analysis.” Journal of Experimental Psychology. General 150 (4): 700–709.

Guyon, Ak ‘ingabe, and Robert Perreault. 2016. “Public Health Systems under Attack in Canada: Evidence on Public Health System Performance Challenges Arbitrary Reform.” Canadian Journal of Public Health. Revue Canadienne de Sante Publique 107 (3): e326–29.

Hellevik, Ottar. 2009. “Linear versus Logistic Regression When the Dependent Variable Is a Dichotomy.” Quality & Quantity 43 (1): 59–74.

Hennessy, Deirdre A., William M. Flanagan, Peter Tanuseputro, Carol Bennett, Meltem Tuna, Jacek Kopec, Michael C. Wolfson, and Douglas G. Manuel. 2015. “The Population Health Model (POHEM): An Overview of Rationale, Methods and Applications.” Population Health Metrics 13 (September):24.

Herrmann, Thomas, William Gleckner, Rania A. Wasfi, Benoît Thierry, Yan Kestens, and Nancy A. Ross. 2019. “A Pan-Canadian Measure of Active Living Environments Using Open Data.” Health Reports / Statistics Canada, Canadian Centre for Health Information = Rapports Sur La Sante / Statistique Canada, Centre Canadien D’information Sur La Sante 30 (5): 16–25.

Hill, M. Anne, and Elizabeth King. 1995. “Women’s Education and Economic Well-Being.” Feminist Economics 1 (2): 21–46.

Hood, Carlyn M., Keith P. Gennuso, Geoffrey R. Swain, and Bridget B. Catlin. 2016. “County Health Rankings: Relationships Between Determinant Factors and Health Outcomes.” American Journal of Preventive Medicine 50 (2): 129–35.

Horsman, John, William Furlong, David Feeny, and George Torrance. 2003. “The Health Utilities Index (HUI): Concepts, Measurement Properties and Applications.” Health and Quality of Life Outcomes 1 (October):54.

Jann, Ben. 2008. “The Blinder–Oaxaca Decomposition for Linear Regression Models.” The Stata Journal 8 (4): 453–79.

Klinenberg, E. 2018. Palaces for the People: How Social Infrastructure Can Help Fight Inequality, Polarization, and the Decline of Civic Life. Penguin Random House.

Lavergne, M. Ruth, and George Kephart. 2012. “Examining Variations in Health within Rural Canada.” Rural and Remote Health 12 (February):1848.

Le Meur, Nolwenn, Fei Gao, and Sahar Bayat. 2015. “Mining Care Trajectories Using Health Administrative Information Systems: The Use of State Sequence Analysis to Assess Disparities in Prenatal Care Consumption.” BMC Health Services Research 15 (May):200.

Luan, Hui, and Daniel Fuller. 2022. “Urban Form in Canada at a Small-Area Level: Quantifying ‘compactness’ and ‘sprawl’ with Bayesian Multivariate Spatial Factor Analysis.” Environment and Planning B: Urban Analytics and City Science 49 (4): 1300–1313.

Malkin, Jennifer, Charles Plante, Navdeep Sandhu, and Cory Neudorf. 2021. “Developing a Profile Survey for Local Public Health Units in Urban Canada: Integrated Knowledge Translation in Practice.” https://uphn.ca/resources/Documents/Publications/Working-Papers/profilektwp.pdf.

McGil Libraries. n.d. “Enhanced Points of Interest (DMTI).” Libraries. Accessed December 6, 2024. https://www.mcgill.ca/library/find/maps/epoi.

Mikkonen, Juha, and Dennis Raphael. 2010. “The Canadian Facts.” York University School of Health Policy and Management.

Missiuna, Sharalynn, Charles Plante, Punam Pahwa, Nazeem Muhajarine, and Cordell Neudorf. 2021. “Trends in Mental Health Inequalities in Urban Canada.” Canadian Journal of Public Health. Revue Canadienne de Sante Publique 112 (4): 629–37.

Murphy, X. Zhang, and C. Dionne. 2010. “Revising Statistics Canada’s Low Income Measure (LIM).” Statistics Canada, Income Statistics Division, June. https://www150.statcan.gc.ca/n1/pub/75f0002m/75f0002m2010004-eng.htm.

Neumark, David. 1988. “Employers’ Discriminatory Behavior and the Estimation of Wage Discrimination.” The Journal of Human Resources 23 (3): 279–95.

Nguena Nguefack, Hermine Lore, M. Gabrielle Pagé, Joel Katz, Manon Choinière, Alain Vanasse, Marc Dorais, Oumar Mallé Samb, and Anaïs Lacasse. 2020. “Trajectory Modelling Techniques Useful to Epidemiological Research: A Comparative Narrative Review of Approaches.” Clinical Epidemiology 12 (October):1205–22.

Oaxaca, Ronald L., and Michael R. Ransom. 1994. “On Discrimination and the Decomposition of Wage Differentials.” Journal of Econometrics 61 (1): 5–21.

Östlin, Piroska, Ted Schrecker, Ritu Sadana, Josiane Bonnefoy, Lucy Gilson, Clyde Hertzman, Michael P. Kelly, et al. 2011. “Priorities for Research on Equity and Health: Towards an Equity-Focused Health Research Agenda.” PLoS Medicine 8 (11): e1001115.

Pampalon, Robert, Denis Hamel, and Philippe Gamache. 2010. “Health Inequalities in Urban and Rural Canada: Comparing Inequalities in Survival according to an Individual and Area-Based Deprivation Index.” Health & Place 16 (2): 416–20.

Park, Hyojun, Anne M. Roubal, Amanda Jovaag, Keith P. Gennuso, and Bridget B. Catlin. 2015. “Relative Contributions of a Set of Health Factors to Selected Health Outcomes.” American Journal of Preventive Medicine 49 (6): 961–69.

Plante, Charles, Thilina Bandara, Lori Baugh Littlejohns, Navdeep Sandhu, Anh Pham, and Cory Neudorf. 2021. “Surveying the Local Public Health Response to COVID-19 in Canada: Study Protocol.” PloS One 16 (11): e0259590.

Plante, Charles, Sharalynn Missiuna, and Cordell Neudorf. 2024. “The Validity and Reliability of Dichotomized Self-Rated Health Under Different Cutpoints.” bioRxiv. 10.1101/2024.04.18.24306035.

Plante, Charles, Sharalynn Missiuna, and Cory Neudorf. 2021. “Urban Income-Related Health Inequalities in Canada: City-Level Results in Health System Use and Self-Reported Indicators.” Urban Public Health Network.

Plante C, Missiuna S and Neudorf C. 2021. “Urban Income-Related Health Inequalities in Canada: City-Level Results in Health System Use and Self-Reported Indicators.” Urban Public Health Network. https://crdcn.ca/publication/urban-income-related-health-inequalities-in-canada-city-level-results-in-health-system-use-and-self-reported-indicators-les-inegalites-en-sante-urbaine-liees-au-revenu-au-canada-resultats-au-nive/.

Potvin, Louise. 2014. “Canadian Public Health under Siege.” Canadian Journal of Public Health. Revue Canadienne de Sante Publique 105 (6): e401–3.

Public Health Agency of Canada (PHAC). 2018. “Key Health Inequalities in Canada: A National Portrait – Executive Summary.” May 15, 2018. https://www.canada.ca/en/public-health/services/publications/science-research-data/key-health-inequalities-canada-national-portrait-executive-summary.html.

Public Health Agency of Canada (PHAC). 2021a. “Expert Advisory Group Report 1: Charting a Path toward Ambition.” June 17, 2021. https://www.canada.ca/en/public-health/corporate/mandate/about-agency/external-advisory-bodies/list/pan-canadian-health-data-strategy-reports-summaries/expert-advisory-group-report-01-charting-path-toward-ambition.html.

Public Health Agency of Canada (PHAC). 2021b. “Expert Advisory Group Report 2: Building Canada’s Health Data Foundation.” Public Health Agency of Canada.

Public Health Agency of Canada (PHAC). 2022. “Expert Advisory Group Report 3: Toward a World-Class Health Data System.” Public Health Agency of Canada. https://www.canada.ca/en/public-health/corporate/mandate/about-agency/external-advisory-bodies/list/pan-canadian-health-data-strategy-reports-summaries/expert-advisory-group-report-03-toward-world-class-health-data-system.html.

Quan, Hude, Vijaya Sundararajan, Patricia Halfon, Andrew Fong, Bernard Burnand, Jean-Christophe Luthi, L. Duncan Saunders, Cynthia A. Beck, Thomas E. Feasby, and William A. Ghali. 2005. “Coding Algorithms for Defining Comorbidities in ICD-9-CM and ICD-10 Administrative Data.” Medical Care 43 (11): 1130–39.

Rao, J. N. K. 2006. “Bootstrap Methods for Analyzing Complex Sample Survey Data.” Statistics Canada.

Reimer, Bill. 2011. “Social Exclusion through Lack of Access to Social Support in Rural Areas.”

Reimer, William. 2004. “Social Exclusion and Social Support in Rural Canada.” In. https://spectrum.library.concordia.ca/id/eprint/6345/.

Schnittker, Jason, and Valerio Bacak. 2014. “The Increasing Predictive Validity of Self-Rated Health.” PloS One 9 (1): e84933.

Sinning, Mathias, Markus Hahn, and Thomas K. Bauer. 2008. “The Blinder–Oaxaca Decomposition for Nonlinear Regression Models.” The Stata Journal 8 (4): 480–92.

Statistics Canada. 2014. “Canadian Classification of the Functions of Government (CCOFOG) Methodology.” November 19, 2014. https://www.statcan.gc.ca/en/statistical-programs/document/5218_D3_T9_V1.

Statistics Canada. 2017a. “Dictionary, Census of Population, 2016.” Ottawa, Ontario: Statistics Canada.

Statistics Canada. 2017b. “Postal Code OM Conversion File.” December 13, 2017. https://www150.statcan.gc.ca/n1/en/catalogue/92-154-X.

Statistics Canada. 2018. “Healthy Behaviours, 2017.” Statistics Canada. 2018. https://www150.statcan.gc.ca/n1/pub/82-625-x/2018001/article/54975-eng.htm.

Statistics Canada. 2019. “Canadian Population Health Survey Data Linked to Mortality, Hospitalization and Historical Postal Codes.” Statistics Canada. June 26, 2019. https://www.statcan.gc.ca/en/microdata/data-centres/data/cphs.

Statistics Canada. 2020. “Census of Population.” May 12, 2020. https://www23.statcan.gc.ca/imdb/p2SV.pl?Function=getSurvey&SDDS=3901.

Statistics Canada. 2021. “2021 Census Boundary Files.” November 17, 2021. https://www12.statcan.gc.ca/census-recensement/2021/geo/sip-pis/boundary-limites/index2021-eng.cfm?year=21.

Statistics Canada. 2023a. “Canadian Classification of Functions of Government (CCOFOG) by Consolidated Government Component (x 1,000,000).” Statistics Canada. November 28, 2023. https://www150.statcan.gc.ca/t1/tbl1/en/tv.action?pid=1010000501.

Statistics Canada. 2023b. “Canadian Community Health Survey - Annual Component (CCHS).” December 18, 2023. https://www23.statcan.gc.ca/imdb/p2SV.pl?Function=getSurvey&SDDS=3226.

Statistics Canada. 2024a. “Canadian Cancer Registry (CCR).” January 30, 2024. https://www23.statcan.gc.ca/imdb/p2SV.pl?Function=getSurvey&SDDS=3207.

Statistics Canada. 2024b. “Canadian Vital Statistics - Death Database (CVSD).” November 19, 2024. https://www23.statcan.gc.ca/imdb/p2SV.pl?Function=getSurvey&SDDS=3233.

Statistics Canada. 2024c. “Census Subdivision Boundary File, 2024.” June 26, 2024. https://www12.statcan.gc.ca/census-recensement/2011/geo/bound-limit/bound-limit-s-eng.cfm?year=24.

Subedi, Rajendra, Nicole Aitken, and Lawson Greenberg. 2002. “Canadian Social Environment Typology User Guide / by Rajendra Subedi, Nicole Aitken, and Lawson Greenberg.: CS172-00002/2022-2E-PDF - Government of Canada Publications - Canada.ca.” July 1, 2002. https://publications.gc.ca/site/eng/9.910159/publication.html.

Statistics Canada. 2022. “Canadian Social Environment Typology User Guide.” Statistics Canada.

Thomas, Steven, and Brenda Wannell. 2009. “Combining Cycles of the Canadian Community Health Survey.” Health Reports / Statistics Canada, Canadian Centre for Health Information = Rapports Sur La Sante / Statistique Canada, Centre Canadien D’information Sur La Sante 20 (1): 53–58.

Vaillant, Nicolas, and François-Charles Wolff. 2012. “On the Reliability of Self-Reported Health: Evidence from Albanian Data.” Journal of Epidemiology and Global Health 2 (2): 83–98.

Weichenthal, Scott, Evi Dons, Kris Y. Hong, Pedro O. Pinheiro, and Filip J. R. Meysman. 2021. “Combining Citizen Science and Deep Learning for Large-Scale Estimation of Outdoor Nitrogen Dioxide Concentrations.” Environmental Research 196 (May):110389.

Wilson, Sarah E., Susan Quach, Shannon E. MacDonald, Monika Naus, Shelley L. Deeks, Natasha S. Crowcroft, Salaheddin M. Mahmud, et al. 2017. “Immunization Information Systems in Canada: Attributes, Functionality, Strengths and Challenges. A Canadian Immunization Research Network Study.” Canadian Journal of Public Health. Revue Canadienne de Sante Publique 107 (6): e575–82.

World Health Organization. 2019. “Healthy, Prosperous Lives for All: The European Health Equity Status Report: Executive Summary.” World Health Organization. https://apps.who.int/iris/bitstream/handle/10665/346044/WHO-EURO-2019-3536-43295-60680-eng.pdf?sequence=3.

World Health Organization (WHO). 2010. “A Conceptual Framework for Action on the Social Determinants of Health.” World Health Organization.

World Health Organization (WHO). 2019. “Healthy, Prosperous Lives for All: The European Health Equity Status Report.” World Health Organization.

Wu, Shunquan, Rui Wang, Yanfang Zhao, Xiuqiang Ma, Meijing Wu, Xiaoyan Yan, and Jia He. 2013. “The Relationship between Self-Rated Health and Objective Health Status: A Population-Based Study.” BMC Public Health 13 (April):320.

Young, Marisa, Sean Leipe, and Diana Singh. 2023. “Best Practices for Measuring Community Resources across Canada: A Comparison of Coding Classifications.” The Canadian Geographer. Geographe Canadien 68 (1): 115–28.

Young, Marisa, and Diana Singh. 2024. “The Canadian Family-Friendly Community Resources Study for Better Balance, Health and Well-Being.” Community, Work & Family 27 (4): 472–98.

